# Diagnostic impact of secondary findings from whole genome or whole exome sequencing in patients diagnosed with genetic eye disorders

**DOI:** 10.1101/2025.03.03.25322102

**Authors:** Setu P. Mehta, Bani Antonio Aguirre, Wendy Y. N. See, Edward P. Esposito, Annabelle Pan, Carolyn Applegate, Kelsey Guthrie, Christy H. Smith, Jefferson J. Doyle, Mandeep S. Singh

## Abstract

**Purpose:** Whole exome or genome sequencing (WES and WGS, respectively) can be valuable in identifying the molecular basis of inherited retinal disorders (IRD), particularly when panel-based testing yields negative or inconclusive results. However, WES or WGS may reveal secondary findings (SF) associated with severe diseases such as cancer, neurodegenerative and cardiovascular conditions. The American Society of Medical Genetics and Genomics has defined a list of reportable SFs that entail significant health implications. Acknowledging the limited awareness of SFs among ophthalmologists, we sought to understand the frequency, characteristics, and impact of SF following WES/WGS.

**Design:** Retrospective cross-sectional study

**Subjects, Participants and/or Controls:** We included patients with suspected IRD who underwent WES or WGS at Johns Hopkins Hospital from 2019-2024. WES/WGS was ordered following genetic counseling and consenting by certified genetic counselors (CGCs).

**Methods/ Main Outcome Measures:** We conducted a qualitative analysis of WES/WGS indications and SF. We also analyzed SF consent status and recommended clinical actions.

**Results:** The study included 36 patients, 50% of whom were females. 34 patients received WES, and two received WGS. The mean age was 45 ± 24 years. 89% (32/36) patients consented to examine SF, while 11% declined analysis. Of the 32 patients who consented to SF analysis, 5 had SFs (16%). Detected SF included pathogenic or likely pathogenic variants in the *APC, RET, TP53, MUTYH,* and *BRCA2* genes associated with familial adenomatous polyposis, multiple endocrine neoplasia, Li-Fraumeni syndrome, *MYH*-associated polyposis, and hereditary breast and ovarian cancer, respectively. After WES/WGS, patients were advised to continue managing their symptoms with their healthcare team, take further medical actions, and/or encourage family members to pursue SF genetic testing. Of the 33 individuals who received post-test genetic counseling, 79% (26/33) were counseled by a CGC, 18% (6/33) by an ophthalmologist, and 3% (1/33) by a primary care provider. IRD gene results were positive in six (16%) patients, none of whom had SF.

**Conclusions:** SF are not infrequent when pursuing WES/WGS for diagnostic testing in IRDs. This study highlights the need for ophthalmologists to be prepared to discuss and manage SF. Multidisciplinary care, including a CGC, enables successful clinical management in this context.

## Introduction

Genetic testing is a critical component of the diagnostic pathway for genetic eye disorders, including inherited retinal disorders (IRDs). Over 270 genes and loci have been identified as causes of IRD^1^; the substantial phenotypic overlap between these genetic subtypes necessitates genetic testing to determine the exact genetic IRD type.

Recognizing the importance of genetic testing, the American Academy of Ophthalmology guidelines recommend genetic testing for all individuals suspected of having an IRD diagnosis^2^. Genetic diagnosis identifies patients who may benefit from gene-specific management. Examples of gene-specific management include voretigene neparvovec-rzyl gene therapy for *RPE65*-associated retinopathy, arginine restriction for gyrate atrophy due to biallelic *OAT* variants, and recommending measures to control vitamin A intake in *ABCA4* retinopathies. Additional benefits of genetic testing include identifying at-risk or affected relatives, enabling family planning, and determining eligibility for clinical trials and research.

Genetic testing modalities encompass single gene testing, gene panels, whole exome sequencing (WES), whole genome sequencing (WGS) and mitochondrial DNA (mtDNA) testing. In general, the diagnostic yield of gene panels is between 40-60% ^1,3,4^. A recent study showed that gene panel testing in retinopathies of unknown origin was nondiagnostic in 41% of cases^5^. In such cases and others with negative or inconclusive gene panel testing, WES/WGS can be used to evaluate additional genes. WES targets all gene exons and some surrounding regions, including parts of introns, while WGS targets the entire genome, including introns, exons, and other regulatory regions. Similar to gene panels, the diagnostic yield of WES and WGS in all diseases is approximately 50-70% ^6–8^.

WES/WGS can produce secondary findings (SF), which are genetic variants associated with conditions unrelated to the primary test indication. The American College of Medical Genetics and Genomics (ACMG) publishes a list of reportable SFs that have significant health implications, including cancer (e.g., breast, gastrointestinal cancers), cardiovascular diseases (e.g., aortic aneurysm, long QT syndrome), endocrine diseases (e.g., pheochromocytoma), neurologic conditions (e.g., Wilson disease), myopathies (e.g., malignant hyperthermia) and others. The current version of this list is ACMG SF v3.2^9,10^.

In light of potential SFs, informed consent for WES/WGS ^4,9,11,12^ is crucial. Prior to testing, patients must consent to testing for serious diseases or risks other than the initial indication. The WES/WGS consent and subsequent disclosure of SF, if detected, entail complex conversations with potentially life-changing implications for the patient and their family. At our institution, these conversations are conducted and documented by certified genetic counselors (CGCs), recognizing that ophthalmologists do not generally possess professional competencies in genetic medicine or genetic counseling.

Although WES/WGS are becoming more integrated into clinical ophthalmology practices, there is a dearth of SF data in the published literature. To our knowledge, there is a single case report of SF arising from WES for an ophthalmic indication of genetic eye disorders^13^. Recognizing this knowledge gap, we aimed to investigate the characteristics, frequency, and diagnostic impact of SF among individuals undergoing WES/WGS for the molecular diagnosis of genetic eye disorders.

## Methods

This retrospective cross-sectional study included participants with a phenotypic diagnosis of a possible genetic eye disorder at the Johns Hopkins Wilmer Eye Institute Genetic Eye Diseases (GEDi) Center who underwent WES or WGS between January 2019 and January 2024. The Johns Hopkins School Medicine Institutional Review Board approved the study, which adhered to the principles of the Declaration of Helsinki. IRD specialists assisted with identifying phenotypic diagnoses using clinical data. Following ophthalmic evaluation, patients were referred to CGCs who counseled for WES/WGS, obtained informed consent, facilitated sample collection, ensured its conveyance to the appropriate Clinical Laboratory Improvement Amendments (CLIA)- certified genetic testing laboratory, received the results, disclosed the results to the patient and ophthalmologist, and coordinated other testing and/or medical referrals as needed.

Data collected included de-identified demographic and clinical information, prior genetic test results, pre- and post-WES/WGS genetic counseling documentation, consent to SF, WES/WGS SF results, and recommended clinical actions after WES/WGS. We include those SFs reported by the testing laboratories, which generally followed ACMG SF guidelines. We obtained written consent for image publication from the three participants whose images are shown in this paper. Descriptive analyses were performed using Microsoft Excel.

## Results

### Study population

A total of 36 participants (50% female) had undergone WES (n=34) or WGS (n=2). The mean age was 45 ± 24 years (range: 4-90 years). Of those, 72% self-identified as White, 11% as Black or African American, 3% as American Indian, 8% as Asian, and 6% as another race (**Table 1**).

**Table 1:** Participant demographics. SD: standard deviation.

| Demographics |  | Frequency |
| --- | --- | --- |
| Total *N* |  | 36 |
| Women |  | 18 (50%) |
| Age (mean ± SD, yrs) | | $45 \pm 24$ |
| Race |  |  |
|  | White | 26 (72%) |
|  | Black or African American | 4 (11%) |
|  | American Indian and Alaska Native | 1 (3%) |
|  | Asian | 3 (8%) |
|  | Other | 2 (6%) |

### Phenotypic diagnosis

Rod-cone dystrophy (i.e., retinitis pigmentosa), the most common diagnosis, accounted for 42% of cases. Macular dystrophy, including Stargardt disease, was diagnosed in 19% of cases. Cone dystrophy and cone-rod dystrophy comprised 11% and 3% of the cases, respectively (**Table 2**). Additionally, 25% of participants had other diagnoses, including foveal hypoplasia, high myopia, and hereditary optic atrophy.

**Table 2:** Phenotypic diagnoses prior to genetic testing. *Other included foveal hypoplasia, high myopia, optic atrophy, blue cone monochromacy and Alport syndrome.

| Diagnosis |  | Frequency | Age Range (years) |
| --- | --- | --- | --- |
| Rod-Cone Dystrophy |  | 15 (42%) | 10-85 |
| Macular Dystrophy |  | 7 (19%) | 25-90 |
|  | *Stargardt Disease* | 2 (6%) | 50-65 |
| Cone Dystrophy |  | 4 (11%) | 10-80 |
| Cone-Rod Dystrophy |  | 1 (3%) | 50-60 |
| Other* |  | 9 (25%) | 0-65 |

### Secondary findings

The decision to pursue WES/WGS was based on negative or inconclusive prior genetic testing (69%), syndromic phenotype (22%), or other reasons (8%) (i.e., patient preference or family history of other diseases).

A total of 4 participants did not consent for SF testing and so SF data were unavailable in those cases. Among those who consented to SF reporting, SF were discovered in 16% (5/32). IRD-causing variants were not found in any participant with SF. WES/WGS was positive for causative eye disease genes in 16% (6/36) of participants.

SFs among participants in this study included gene variants associated with attenuated familial adenomatous polyposis (*APC*), multiple endocrine neoplasia (*RET*), Li-Fraumeni syndrome (*TP53*), colorectal cancer (*MUTYH)* as well as hereditary breast or ovarian cancer (*BRCA2*). **Table 3** lists the ages, phenotypic diagnoses, genetic variants, and clinical relevance of SFs. Post-SF recommendations included additional SF-related specialist evaluation for the proband and testing family members for the SF-related condition.

**Table 3:** List of secondary findings (n=5). None of these patients had pathogenic IRD variants identified through WES/WGS. *Biallelic *MUTYH* variants are associated with polyposis, adenoma, and multiple colorectal and gastric cancers; however, this patient had a single variant that was interpreted as conferring a marginally increased or unclear risk of cancer. AD: autosomal dominant; AR: autosomal recessive. Age ranges (0-18, 19-36, 37-54, 55-72, >73) are shown instead of actual age to minimize potential identifiers.

| Age Range | Phenotypic Diagnosis | Test | Secondary Finding | Inheritance | Clinical Relevance |
| --- | --- | --- | --- | --- | --- |
| 0-18 | Rod-cone dystrophy | WGS | *MUTYH* c.462+1G>T | AR | Polyposis, adenoma, and multiple colorectal and gastric cancers* |
| 0-18 | Optic atrophy | WES | *BRCA2* c.5350_5351del | AD | Hereditary breast and ovarian cancer |
| 37-54 | Macular dystrophy | WES | *APC* c.5065dup | AD | Attenuated familial adenomatous polyposis |
| 37-54 | Cone-rod dystrophy | WES | *RET* c.2410 G>A | AD | Multiple endocrine neoplasia type 2 and familial medullary thyroid cancer |
| 55-72 | Rod-cone dystrophy | WES | *TP53* c.817C>T | AD | Li-Fraumeni syndrome |

### Genetic counseling

Prior to genetic testing, all patients in this study met with a CGC for pre-test genetic counseling and consent. The documented discussion included indications and rationale for genetic testing, the benefits, risks, and limitations of WES/WGS, the potential for SFs and implications for systemic health, and specific consent for SF testing.

After WES/WGS results were obtained, all patients were notified of the results and 92% of those had documentation of post-WES/WGS genetic counseling. A CGC counseled 79% (26/33) of these patients, an ophthalmologist counseled 18% (6/33), and another healthcare worker counseled 3% (1/33). Participants with SF were recommended to complete one or more of the following actions: continue to manage their ongoing symptoms with their healthcare team, take further medical action such as scheduling additional medical appointments, and recommend genetic testing to family members.

### Illustrative Cases

***Case 1.*** A patient aged between 37-54 was diagnosed clinically with macular pattern dystrophy (**Figure 1**). Best corrected visual acuity was 20/100 in the right eye (OD) and 20/63 in the left eye (OS). A previous IRD gene panel (My Retina Tracker Program Panel Plus, 266 genes) was negative. WES was negative for IRD genes. However, a likely pathogenic variant in *APC* (c.5065dup, p.T1689Nfs*11) was detected. Due to the associated risk of attenuated familial adenomatous polyposis and the 70% lifetime risk of colorectal cancer (CRC)^14^, the CGC arranged oncology and colonoscopy referrals, and CGC helped to facilitate conversations with the participant’s family about the genetic testing for *APC*.

**Figure 1.**
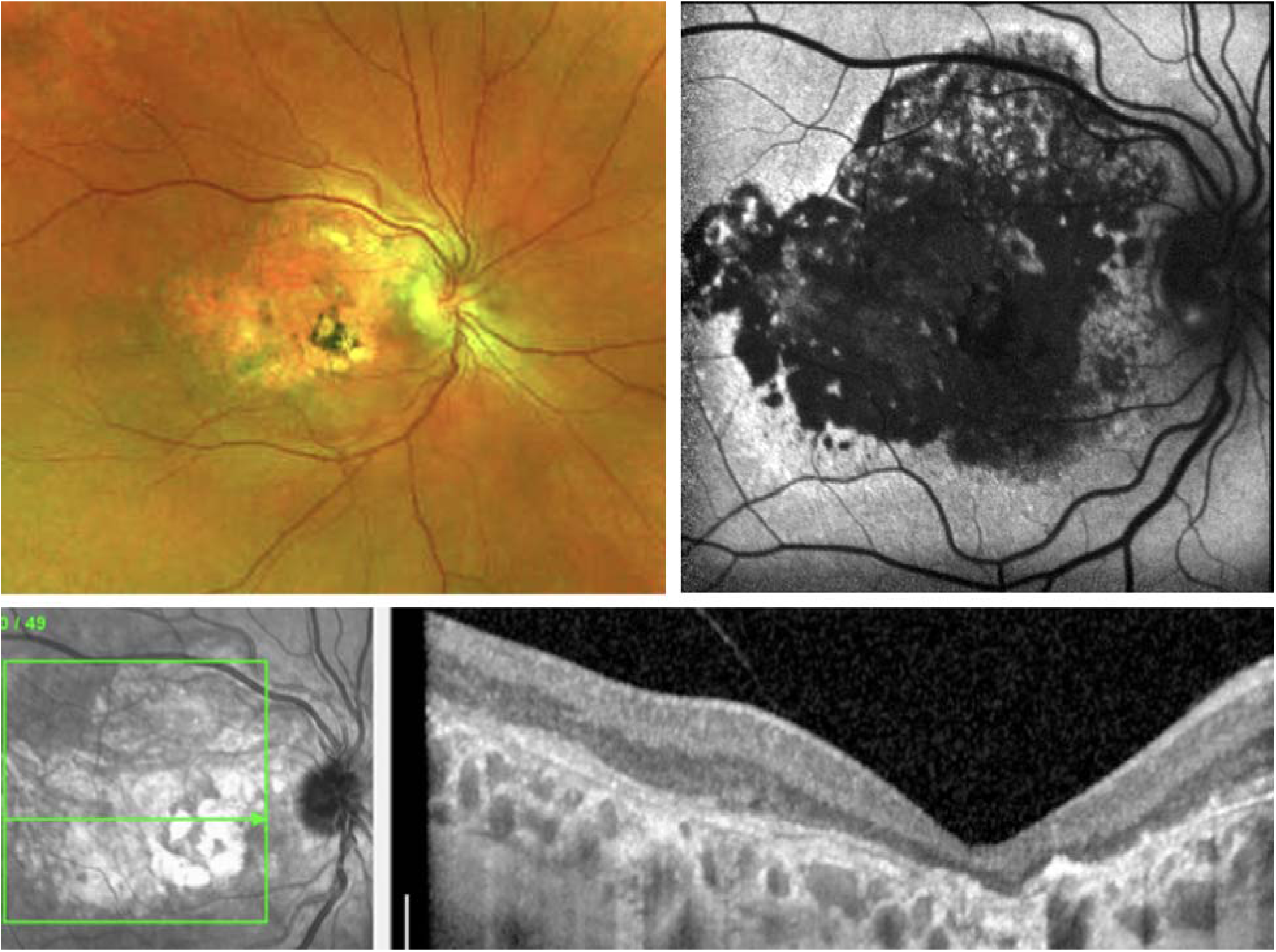
In the right eye of case 1, pseudocolor fundus photography (**top left**) showed a macular atrophy and hyperpigmentary changes, consistent with a phenotypic diagnosis of macular dystrophy. Visual acuity was 20/100 in the right eye (OD) and 20/63 in the left eye (OS). Fundus autofluorescence imaging (**top right**) showed confluent areas of hypoautofluorescence and no hyperautofluorescent flecks. Optical coherence tomography (**bottom**) showed outer nuclear layer degeneration and subretinal fibrosis. The findings in the left eye were qualitatively similar.

***Case 2.*** A patient aged between 0 to 18 presented with retinitis pigmentosa (RP) (**Figure 2**) with BCVA 20/30 OD and 20/25 OS. A previous IRD gene panel (MyRetina Tracker Panel with mitochondrial genes, 351 genes) was negative. WGS did not identify a genetic cause of RP. However, a likely pathogenic variant in the *MUTYH* gene (c.462+1G>T, p.-), associated with adenomas and colorectal cancer, was identified. It is important to note that heterozygosity for this *MUTYH* variant confers only a marginal risk of CRC and an unclear risk for extraintestinal cancer^15^. This patient was referred to oncology, and the patient’s parents met with a prenatal geneticist to discuss subsequent reproductive risks and options.

**Figure 2:**
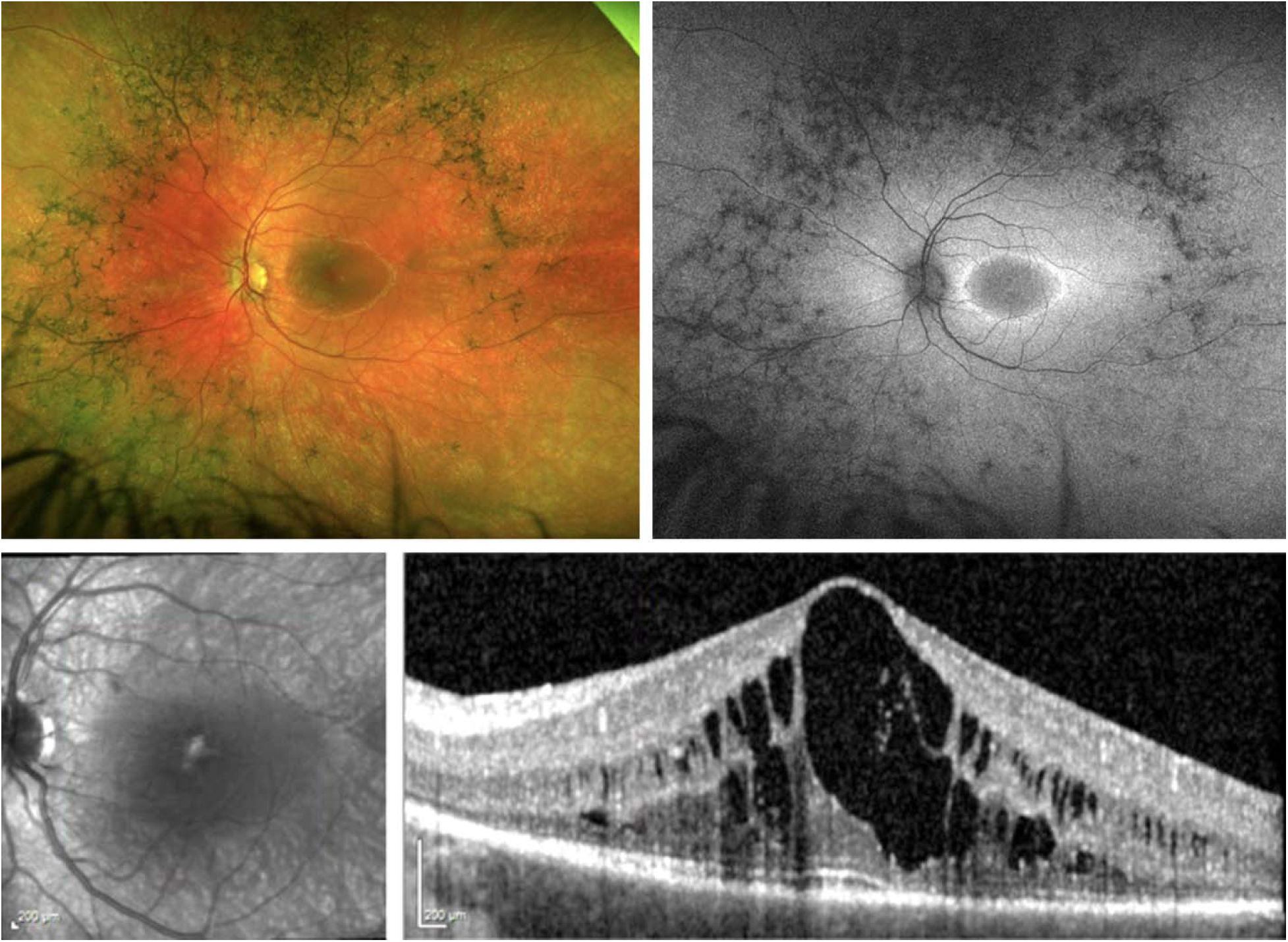
In the left eye of case 2, pseudocolor fundus photography (**top left**) showed a midperipheral bone spicule pigmentation and diffuse vascular attenuation, consistent with a phenotypic diagnosis of retinitis pigmentosa. Visual acuity was 20/30 OD and 20/25 OS. Fundus autofluorescence imaging (**top right**) showed a hyperautofluorescent macular ring and midperipheral bone spicule hypoautofluorescence. Optical coherence tomography (**bottom**) showed outer nuclear layer degeneration, ellipsoid zone attenuation and cystoid macular edema. The findings in the right eye were qualitatively similar.

***Case 3.*** A patient aged between 37-54 was diagnosed with progressive rod-cone dystrophy with light perception (LP) VA in both eyes (**Figure 3**). A previous IRD gene panel (Molecular Vision NGS Retinal Dystrophy Panel, 280 genes) was inconclusive. WES did not identify a genetic etiology for rod-cone dystrophy but revealed an autosomal dominant *RET* gene variant (c.2410 G>A, p.V804M) associated with multiple endocrine neoplasia type 2 (MEN2) or familial medullary thyroid cancer (MTC).

**Figure 3:**
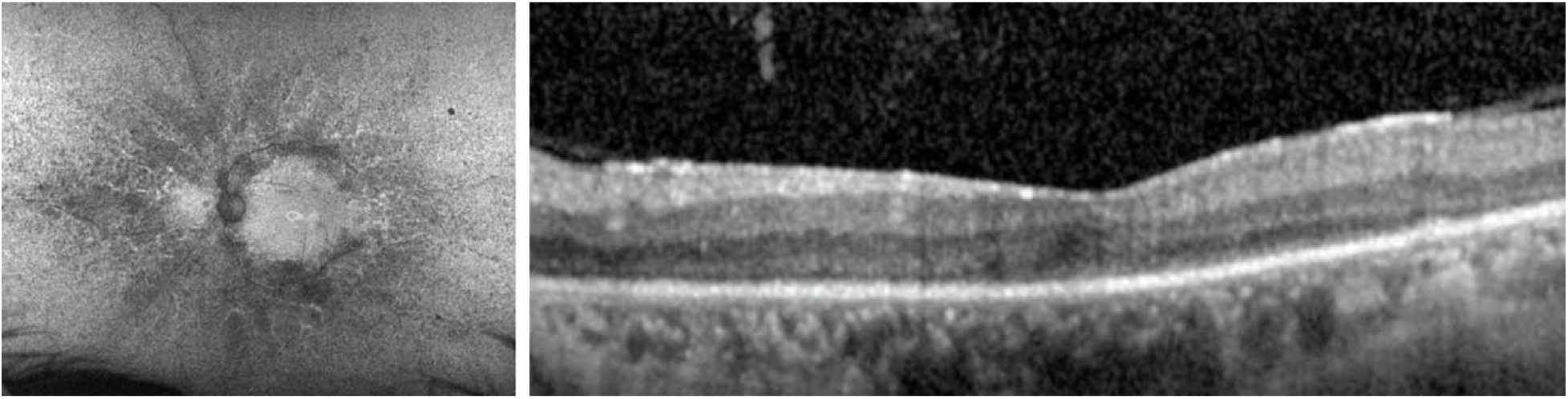
In the left eye of case 3, fundus autofluorescence imaging (**left**) showed a small hyperautofluorescent macular ring with pericentric and midperipheral spicule hypoautofluorescence, consistent with a phenotypic diagnosis of retinitis pigmentosa. Visual acuity was light perception in both eyes. Optical coherence tomography (**right**) showed outer nuclear layer degeneration, ellipsoid zone loss and mild epiretinal membrane. Pseudocolor fundus photography was not obtained. The findings in the right eye were qualitatively similar.

Consequently, the patient was referred to an endocrinologist and was found to have an elevated serum calcitonin level consistent with MEN2 syndrome and early medullary thyroid carcinoma. The patient underwent a prophylactic total thyroidectomy. Testing of relatives was recommended, and the patient reported that, as a result, one of their family members was diagnosed with MTC and underwent cancer treatment.

## Discussion

The data show that SF were found in almost 1/5^th^ of participants in this study who consented to SF – a frequency that generally exceeds the reported SF rates in the context of genetic testing for other (non-IRD) conditions (1–6%)^11^. Broadly, these data underscore the importance of clinician awareness of the possibility of WES/WGS SF and ensuring a robust informed consent process ideally led by a CGC or genetic medicine specialist.

Our SF detection frequency may have been relatively high for several reasons, including a small sample size. Further, our study occurred at an academic medical center that may be enriched with patients with more complex conditions who harbor additional genetic diseases as compared to other healthcare settings. Additionally, the ACMG SF list has grown recently and this may have led to a higher SF detection rate than prior studies.^12^

The diagnostic IRD yield of WES/WGS in this study was 16%, considerably lower than reported yields in other contexts^3,6,7^. It is important to note that the yield of WES/WGS may be lower among cases with a previous negative panel in comparison to using WES/WGS as the initial genetic test^16^.

WES/WGS testing laboratories may follow different conventions to report unsolicited findings (UF). UFs are genetic predispositions (including but not limited to SF) for diseases that exceed the initial diagnostic question. Several laboratories follow the ACMG SF list, but other laboratories may report other UFs that may or may not be medically actionable. Generally, only a minority of labs report UFs related to neurodegenerative diseases. While the ACMG SF list emphasizes medically actionable findings, a healthcare provider has no legal obligation to disclose or act upon SF.

Robust CGC-led consent discussions may positively contribute to patient understanding and agreement to SF testing^17^. The informed consent discussion for WES/WGS in our study was conducted by the CGC and included reviewing the basic principles of genetics, the limitations and advantages of genetic testing, and the possible outcomes. These conversations also emphasized the possibility of SF, the possibility of identifying treatable or non-treatable diseases, the risk to relatives (including children), and the impact on family planning. The latter part of the discussion was relevant to our study as two of the six individuals with SF were younger than 18. Asymptomatic or unaffected children are generally not tested for conditions that manifest in adulthood and conditions that lack medical management options in childhood. In this context, providers may wait to test children for such genetic conditions until adulthood ^17,18,19^.

Patients may decline to test SF for several reasons. For example, patients may fear the physical or mental health impacts of SF. In addition, patients may be concerned about their personal or family health privacy in the context of potential genetic discrimination. CGCs can help by discussing the Genetic Information Nondiscrimination Act of 2008, which protects the privacy of patients’ genetic results^20,21^.

Saelaert et al. (2021) studied how IRD patients may react to UF. First, patients were concerned about the medical implications of UFs, such as how actionable the finding was and the expected age of onset of the anticipated UF disease. Second, patients were concerned about the impact of UFs on the course of their IRD. Lastly, patients were concerned about the impact of UFs on their families^22,23^. Overall, it can be complex for non-qualified providers to assist patients in understanding and acting upon SFs to reduce future morbidity and mortality^9^ because factors such as the patient’s age, current symptoms, and family dynamics may pose decision-making and communication challenges. It is also important to appreciate the potential anxiety that patients may have about sharing SF or UF information with family members. This may hamper expedient communication and thus delay family members’ health evaluations^22^.

A case study reported by Zhu et al. (2022) illustrates the substantial impact of SF on family members. A 23-year-old female with RP underwent trio WES (i.e., testing of the proband and both biological parents) which can increase pathogenic variants detection rate and decrease the inconclusive finding rate^24,25^. A pathogenic variant in the *PMS2* gene (c.137 G>T, p.Ser46Ile), associated with Lynch syndrome and hereditary non-polyposis, was detected in the proband and her mother. Both the patient and her mother were informed of the results, risks to immediate and extended relatives, and management options, including prophylactic surgery. ^13^.

In our study, about 1 in 5 participants with SF received post-test counseling from their ophthalmologist. Most patients will continue seeing ophthalmologists for long-term IRD management; some may be geographically isolated from a CGC. Thus, ophthalmologists who lack CGC support should be comfortable interpreting SF results, facilitating discussions about SF, and managing SF, including referring to other specialists as appropriate^20^. In such cases, it could be helpful for ophthalmologists to partner virtually with a CGC to enable comprehensive care delivery ^26^.

Our study was limited by its small size and retrospective nature. We relied upon documentation from our institution’s CGCs to identify participants. It is possible that some WES/WGS cases did not see the CGC and, thus, were not included in our study. Furthermore, the health records may not have included the full extent of conversations between patients, providers, and relatives. Lastly, our population mainly consisted of patients from White/European backgrounds. Minority populations remain underrepresented in genetic research, and it is vital to support access to and counseling for WES and WGS in these populations^27^.

As many of our patients remained molecularly undiagnosed despite undergoing WES/WGS, our project illustrates the real-world challenges in identifying a genetic cause for IRDs. Further, WES/WGS may have limitations in identifying pathogenic variants in non-coding or repetitive regions ^28,29^. These considerations are critical in cases of X-linked RP, blue cone monochromacy, and Stargardt disease, for example. Therefore, the informed consent process must ensure patients understand that WES/WGS may be non-diagnostic for the IRD indication.

In conclusion, our study showed that SF were not infrequent when pursuing WES/WGS. This highlights the need for ophthalmologists to understand the implications of ordering WES/WGS beyond identifying a genetic etiology for a patient’s genetic eye disease. Multidisciplinary care, including a CGC, is critical to successfully managing patients in this context.

## Data Availability

All data produced in the present study are available upon reasonable request to the authors.

## Acknowledgments

Funding/ Support: Foundation Fighting Blindness CD-RM-0918-0749-JHU (MSS), Joseph Albert Hekimian Fund 1706611301 (MSS), Andreas C. Dracopoulos Professorship (MSS), Dracopoulos-Finkelstein Rising Professorship (JJD)

a. Financial Disclosures: None
b. Other Acknowledgments: None

